## Supplementary information for "Imputation strategies for missing baseline neurological assessment covariates after traumatic brain injury: A CENTER-TBI study"

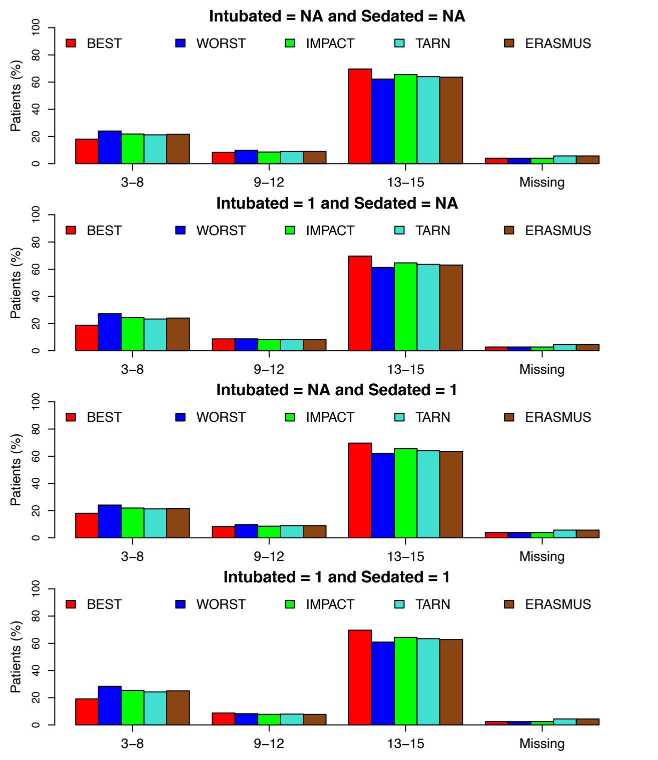


**Figure S1.** GCS distribution between 'mild', 'moderate' and 'severe' categories for all strata

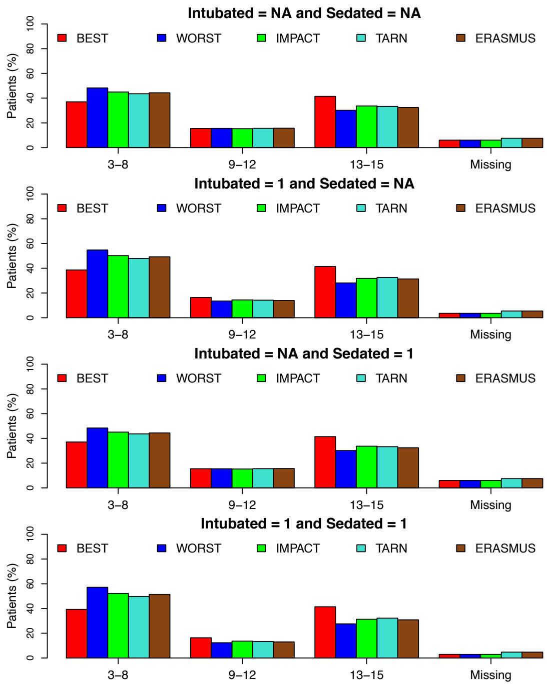


**Figure S2.** GCS distribution between 'mild', 'moderate' and 'severe' categories for the ICU stratum

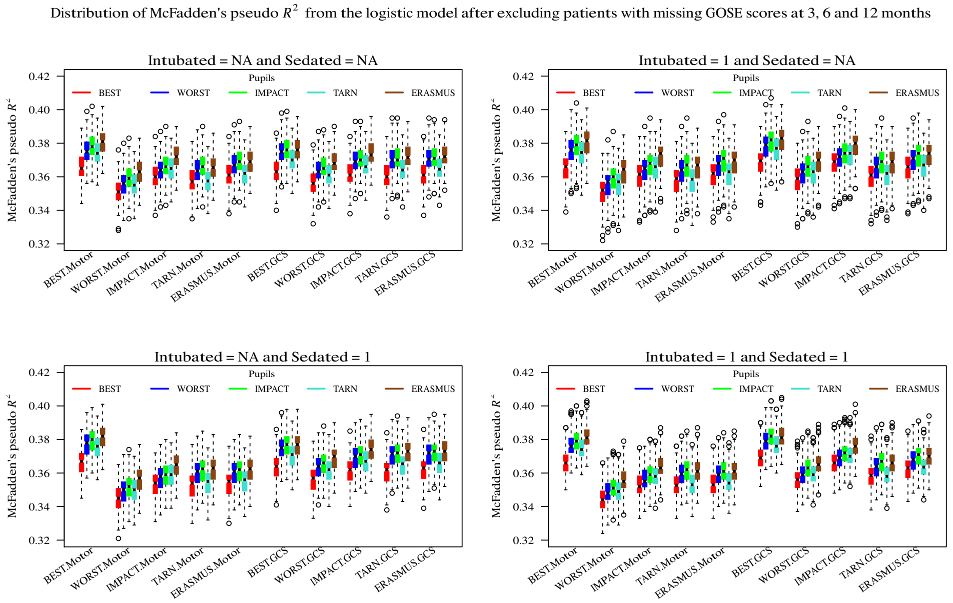


**Figure S3.** Comparison of logistic regressions for dichotomous 6 month survival/death for different combinations of GCS, GCS-motor score and pupil response imputation choices. The boxes/whiskers reflect the variability from the 200 imputed data sets used. Data shown the ICU stratum.

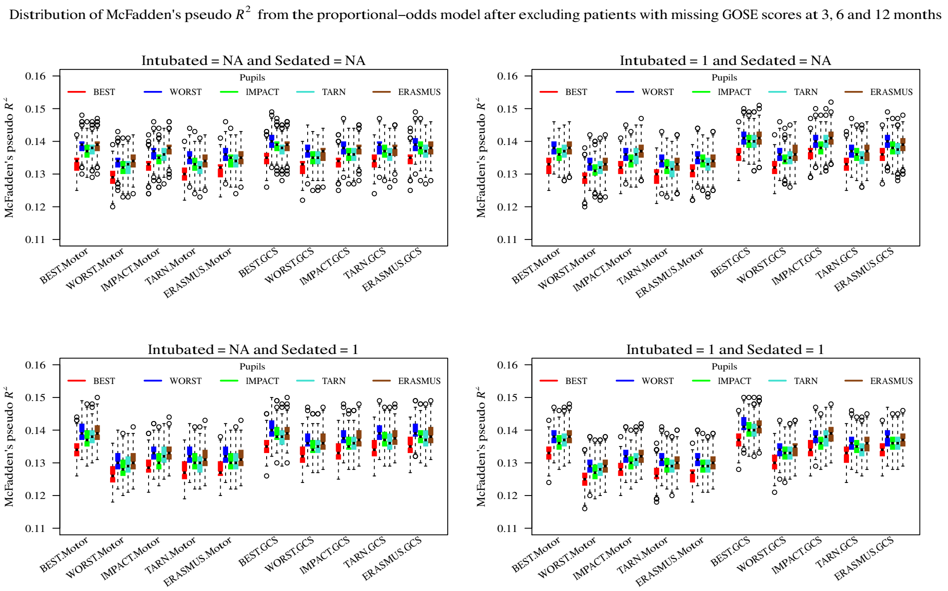


**Figure S4.** Comparison of pooled proportional odd regressions for GOSE for different combinations of GCS, GCS-motor score and pupil response imputation choices. The explanatory value of the prehospital time point is consistently limited. The boxes/whiskers reflect the variability from the 200 imputed data sets used. Data shown for ICU stratum.

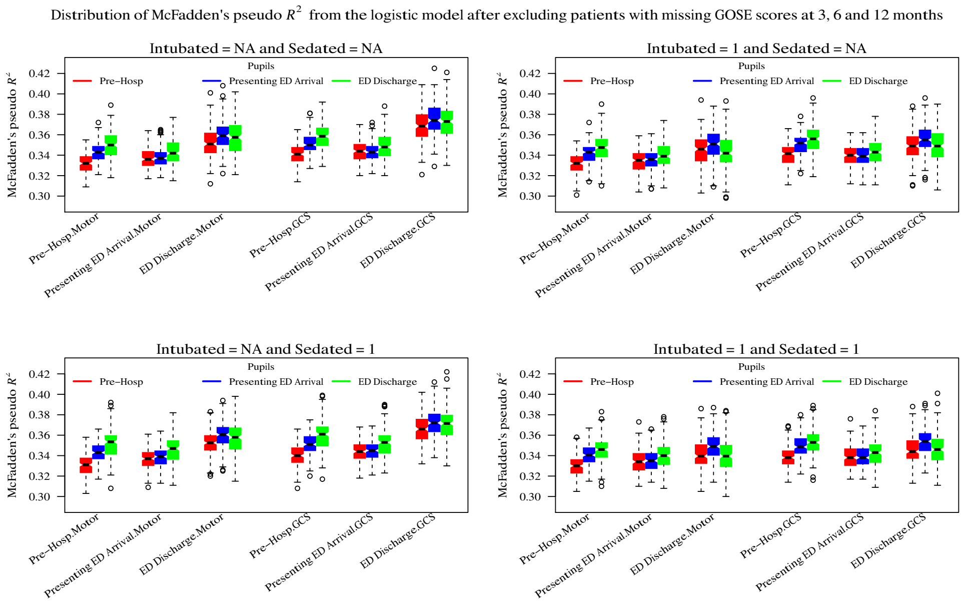


**Figure 5.** Comparison of pooled logistic regressions for 6-month GOSE for fully imputed timepoints. The 'presenting' ED arrival time point was a composite formed from the 'referring' and 'study hospital' ED time points to reflect the first contact with the ED irrespective of whether the patient underwent secondary transfer or not. The boxes/whiskers reflect the variability from the 200 imputed data sets used. Data shown for the ICU stratum.

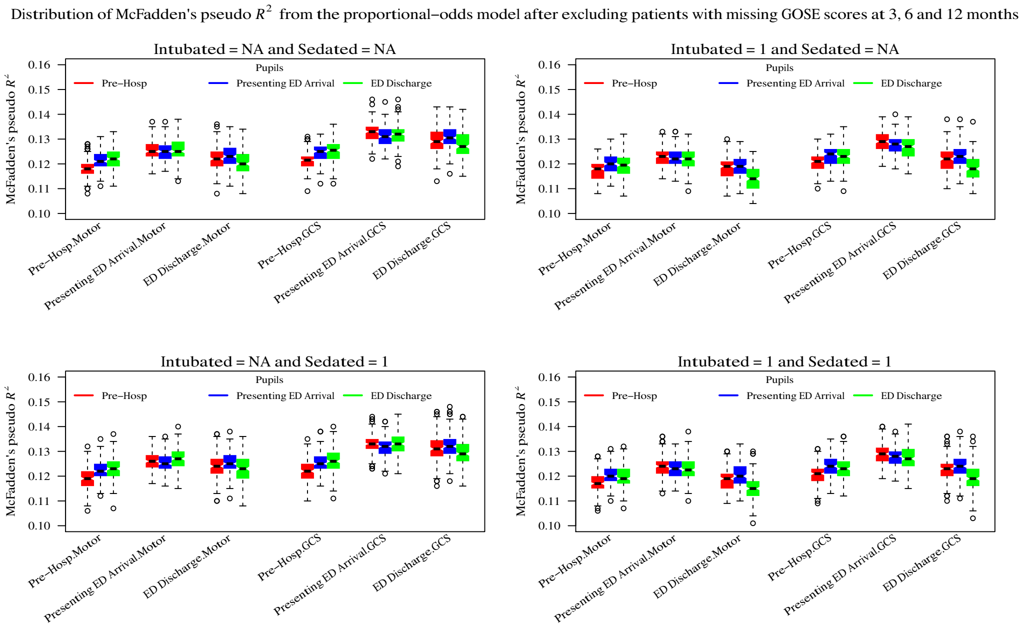


**Figure 6.** Comparison of pooled proportional odds regressions for 6-month GOSE for fully imputed time points. The 'presenting' ED arrival time point was a composite formed from the 'referring' and 'study hospital' ED time points to reflect the first contact with the ED irrespective of whether the patient underwent secondary transfer or not. The boxes/whiskers reflect the variability from the 200 imputed data sets used. Data shown for the ICU stratum
